# Characterization of the emerging B.1.621 variant of interest of SARS-CoV-2

**DOI:** 10.1101/2021.05.08.21256619

**Authors:** Katherine Laiton-Donato, Carlos Franco-Muñoz, Diego A. Álvarez-Díaz, Hector Alejandro Ruiz-Moreno, José A. Usme-Ciro, Diego Andrés Prada, Jhonnatan Reales-González, Sheryll Corchuelo, María T. Herrera-Sepúlveda, Julian Naizaque, Gerardo Santamaría, Jorge Rivera, Paola Rojas, Juan Hernández Ortiz, Andrés Cardona, Diana Malo, Franklin Prieto-Alvarado, Fernando Ruiz Gómez, Magdalena Wiesner, Martha Lucia Ospina Martínez, Marcela Mercado-Reyes

## Abstract

The genetic diversity of severe acute respiratory syndrome coronavirus 2 (SARS-CoV-2) has the potential to impact the virus transmissibility and the escape from natural infection- or vaccine-elicited neutralizing antibodies. Here, representative samples from circulating SARS-CoV-2 in Colombia between January and April 2021, were processed for genome sequencing and lineage determination following the nanopore amplicon ARTIC network protocol and PANGOLIN pipeline. This strategy allowed us to identify the emergence of the B.1.621 lineage, considered a variant of interest (VOI) with the accumulation of several substitutions affecting the Spike protein, including the amino acid changes T95I, Y144T, Y145S and the insertion 146N in the N-terminal domain, R346K, E484K and N501Y in the Receptor-binding Domain (RBD) and P681H^1^ in the S1/S2 cleavage site of the Spike protein. The rapid increase in frequency and fixation in a relatively short time in Magdalena, Atlántico, Bolivar, Bogotá D.C, and Santander that were near the theoretical herd immunity suggests an epidemiologic impact. Further studies will be required to assess the biological and epidemiologic roles of the substitution pattern found in the B.1.621 lineage.

**Highlights:**

- Monitoring the emergence of new variants of SARS-CoV-2 in real time is a worldwide priority.
- Emerging variants of SARS-CoV-2 may have high impact biological implications for public health
- The SARS-CoV-2 B.1.621 variant of interest was characterized by several substitutions: T95I, Y144T, Y145S, ins146N, R346K, E484K, N501Y and P681H in spike protein.

## 1. Introduction

In September 2020, SARS-CoV-2 variants of concern (VOC) and variants of interest (VOI) started to be reported, with more distinctive substitutions than expected from the characteristic clock-like molecular evolution of this virus evidenced during the first year pandemic (Abdool Karim and de Oliveira, 2021; ECDE, 2021). In the following months, with data from clinical and genomic surveillance worldwide, the US government interagency group and Centers for Disease Control and Prevention (CDC) proposed a hierarchical variant classification scheme with three classes of SARS-CoV-2 variants where the higher classes include the possible attributes of lower classes; 1) Variant of Interest, 2) Variant of Concern and 3) Variant of High Consequence (VOHC). A VOI is characterized by a set of Spike protein substitutions associated with increased infectivity, resistance to post-vaccinal/infection antibodies, possible increase in transmissibility and worse clinical outcome. VOC, besides the possible attributes of VOI, there is evidence of impact on diagnostics, treatments, or vaccines, increased transmissibility, and disease severity. VOHC, although currently there are no SARS-CoV-2 identified within this class to date, it is expected that besides the possible attributes of a VOC, a VOHC has strong evidence of diagnostic failure, a significant reduction in vaccine effectiveness, approved therapeutics, and more severe clinical disease and increased hospitalizations (Centers for Disease Control and Prevention, 2021).

Despite mutations spanning the whole genome, an interesting feature of these emerging variants has been the presence of several amino acid substitutions falling in the Spike protein, the viral protein responsible for receptor binding and membrane fusion and also the main target for neutralizing antibodies (NAb) (Greaney et al., 2021). Monitoring the emergence of new variants of SARS-CoV-2 is a priority worldwide, as the presence of certain non-synonymous substitutions and Insertion–deletion mutations (INDELs) could be related to biological properties, such as altering the ligand-receptor affinity, the efficiency of neutralization by naturally acquired polyclonal immunity or post-vaccination antibodies and transmission capacity (Davies et al., 2021; Jeyanathan et al., 2020; Rees-Spear et al., 2021).

In Colombia, the National Genomic Characterization Program led by the Instituto Nacional de Salud has carried out real-time monitoring of the SARS-CoV-2 lineages since the beginning of the pandemic through next generation sequencing and following the Pan American Health Organization (PAHO) guidance for SARS-CoV-2 samples selection for genomic characterization and surveillance (INS, 2021; Laiton-Donato et al., 2020). Until December 2020, over thirty lineages were circulating inside the country without evidence of VOC and VOI importation. However, a lineage turnover accompanied the third epidemic peak during March and April 2021, involving the emergence of B.1 lineage descendants with high mutation accumulation (B.1.621 and the provisionally assigned B.1+L249S+E484K) (PAHO, n.d.), as well as the introduction of the B.1.1.7, P.1 and VOI in Magdalena, Atlántico, Bolivar, Bogotá D.C, and Santander.

In this study, we reported the emergence and spread of the novel B.1.621 lineage of SARS-CoV-2, a new VOI with the insertion 146N and several amino acid substitutions in the Spike protein (T95I,Y144T, Y145S, R346K, E484K, N501Y and P681H).

## 2. Materials and methods

A total of 471 nasopharyngeal swab specimens from patients with positive real time RT-PCR for SARS-CoV-2, were collected between January 1st and April 30th, 2021were selected from routine surveillance in all departments based on the representativeness and virologic criteria (PAHO, n.d.). Samples were processed by using the amplicon sequencing protocol v3 (https://www.protocols.io/view/ncov-2019-sequencing-protocol-v3-locost-bh42j8ye). The assembly of raw NGS data was performed by following the pipeline described for Oxford Nanopore Technologies (ONT) platform (https://artic.network/ncov-2019/ncov2019-bioinformatics-sop.html).

Lineage assignment started by filing a new issue in the pango-designation repository (https://github.com/cov-lineages/pango-designation/issues/57) followed by designation as B.1.621 lineage by the Pangolin curation team and PangoLEARN model training for subsequent automatic lineage assignment.

Dataset was aligned using the MAFFT v.7 software and maximum likelihood tree reconstruction was performed with the GTR+F+I+G4 nucleotide substitution model using IQTREE. Branch support was estimated with an SH-like approximate likelihood ratio test (SH-aLRT) and ultrafast bootstrap. Recombination detection was performed using RDP4 software with RDP, GENECONV, Bottscan, Maxchi, Chimaera, SiSscan, and 3Seq tests (*P*-value□<□0.05). Dataset 1 included Colombian SARS-CoV-2 sequences representative of the different lineages and dataset 2 included sequences previously reported as VOC or VOI. Adaptive evolution analysis at the codon level was estimated by Hyphy using stochastic evolutionary models. The detection of individual sites was performed with methods such as MEME (Mixed Effects Model of Evolution), and FEL (Fixed Effects Likelihood) (*P*-value <0.3).

## 3. Results

The routine genomic surveillance of SARS-CoV-2 in Colombia was reinforced in January 2021 for a higher sensitivity monitoring of the potential importation of VOC. By May 7, 2021, a total of 908 sequences from Colombia were available in the GISAID database. Lineage B.1 is the best-represented lineage (with 229 records) due to its higher frequency from the beginning of the pandemic. The recently designated B.1.621 lineage has been increasingly detected since January 11, 2021 (collection date of the first genome belonging to the lineage) to date (77 records), occupying the fifth place in frequency (Figure 1A), and rapidly becoming fixed in some departments located in the North of the country or co-circulating with other lineages in Bogotá D.C. and Santander (Figure 1B).

**Figure 1.**
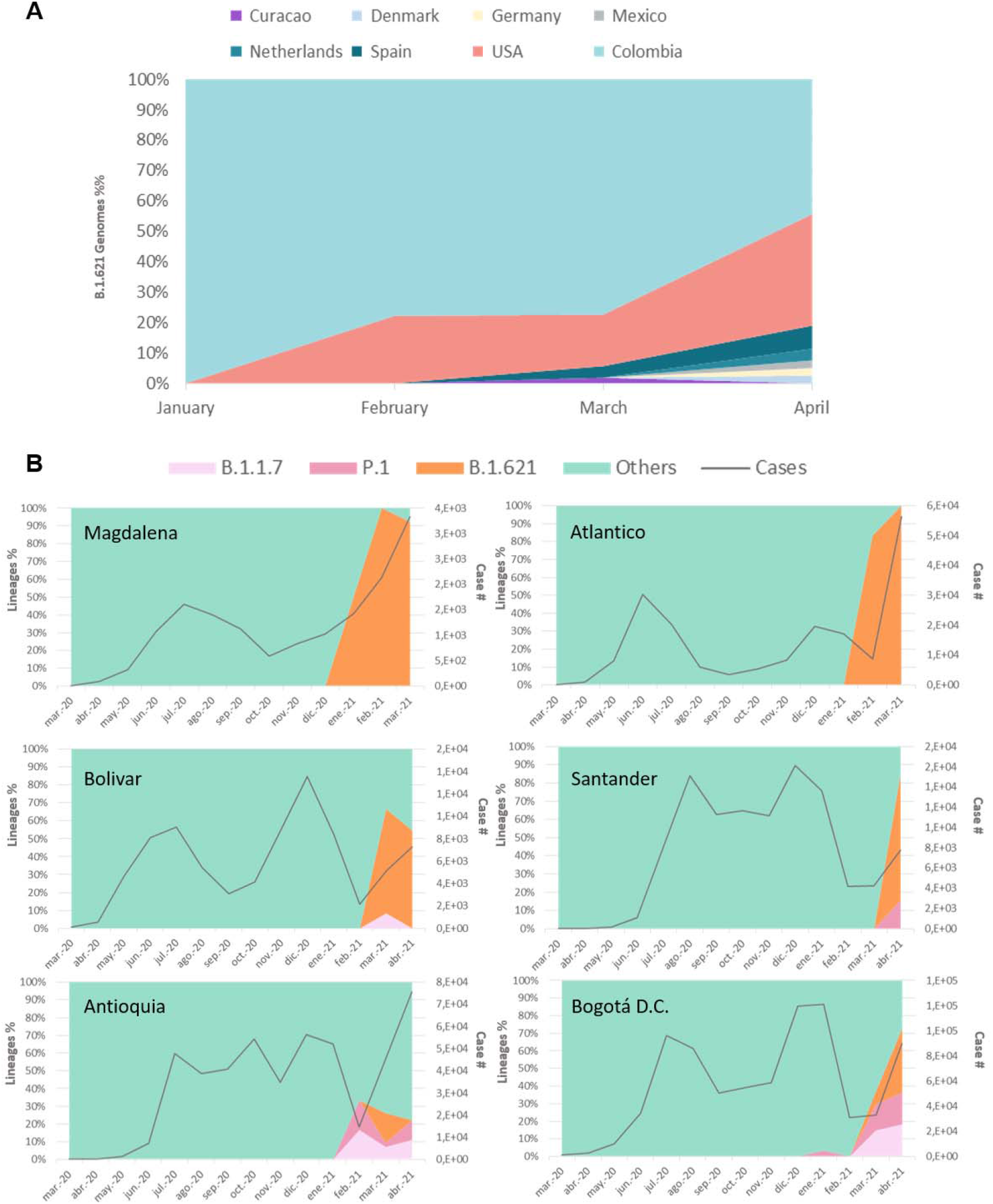
Percent of variant B.1.621 in Spain, USA and Colombia.1A) Gisaid registries of variant B.1.621 in 2021. Since January the continuous record has been maintained in Colombia. 1B) Lineage percentage and number of cases of COVID-19 in five departments with circulation of B.1.621 variant and the capital city. B.1.1.7 and P.1 VOC lineages are shown, others lineages circulating are represented as “others”.

The original assignment through the Pangolin algorithm for this monophyletic group was the B.1 lineage. The genetic background of the B.1.621 lineage includes some convergent amino acid changes in the RBD of the Spike protein which have appeared independently in several VOI and VOC: N501Y, present in B.1.1.7, B.1.351, and P.1 lineages; and E484K present in B.1.351 and P.1 (Harvey et al., 2021) as well as P681H in the S1/S2 furin cleavage site in B.1.1.7 and B.1.1.50 + P681H Variant (Zuckerman et al., 2021). However, a distinctive profile of synonymous and non-synonymous substitutions was found in the Spike protein of B.1.621, including T95I, Y144T, Y145S in the N-terminal domain in addition to substitutions R346K, E484K and N501Y in the RBD, P681H in the S1/S2 furin cleavage site (Table 1) and the insertion 146N in the N-terminal domain (NTD) (supplementary table 1).

**Table1.**
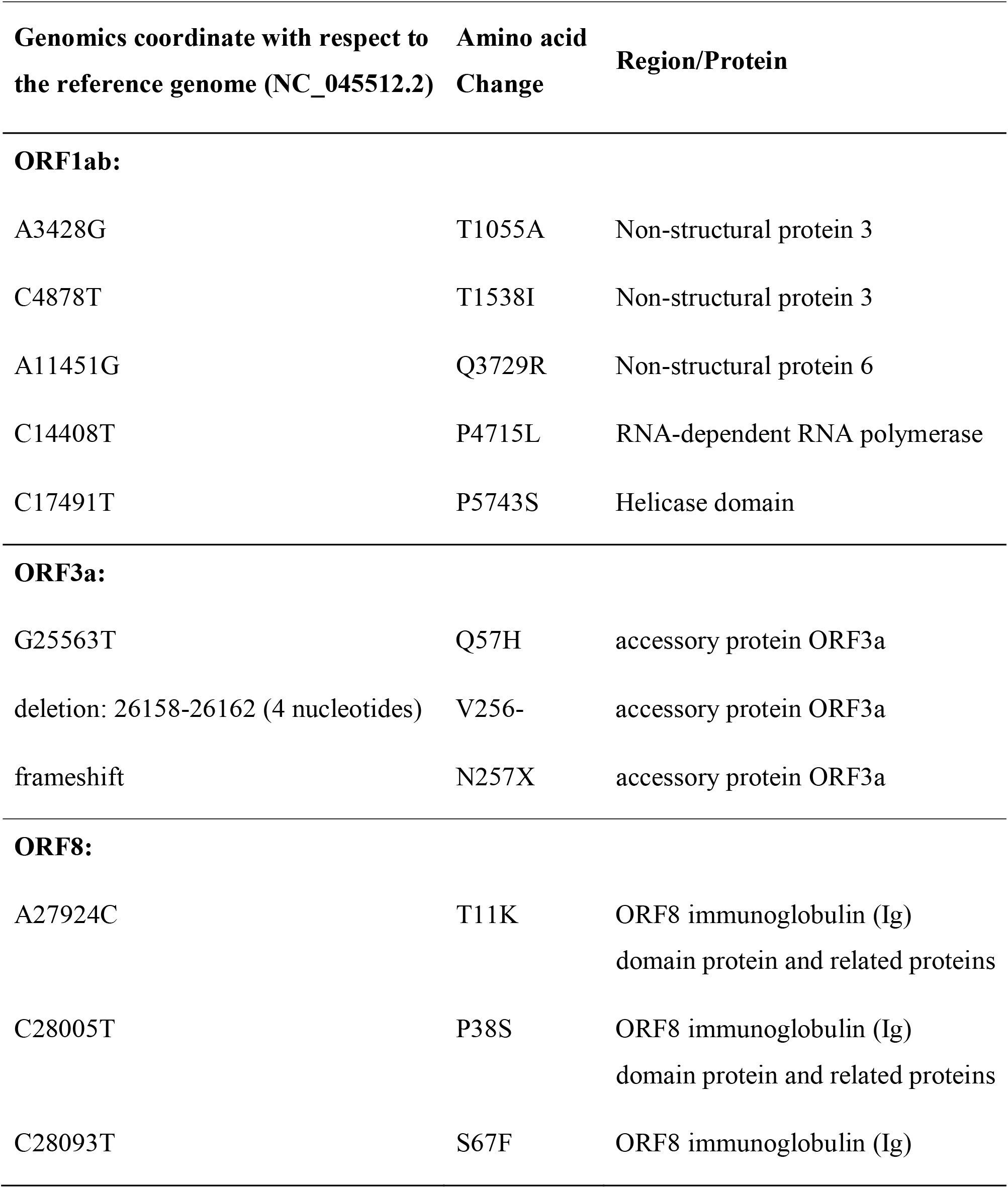
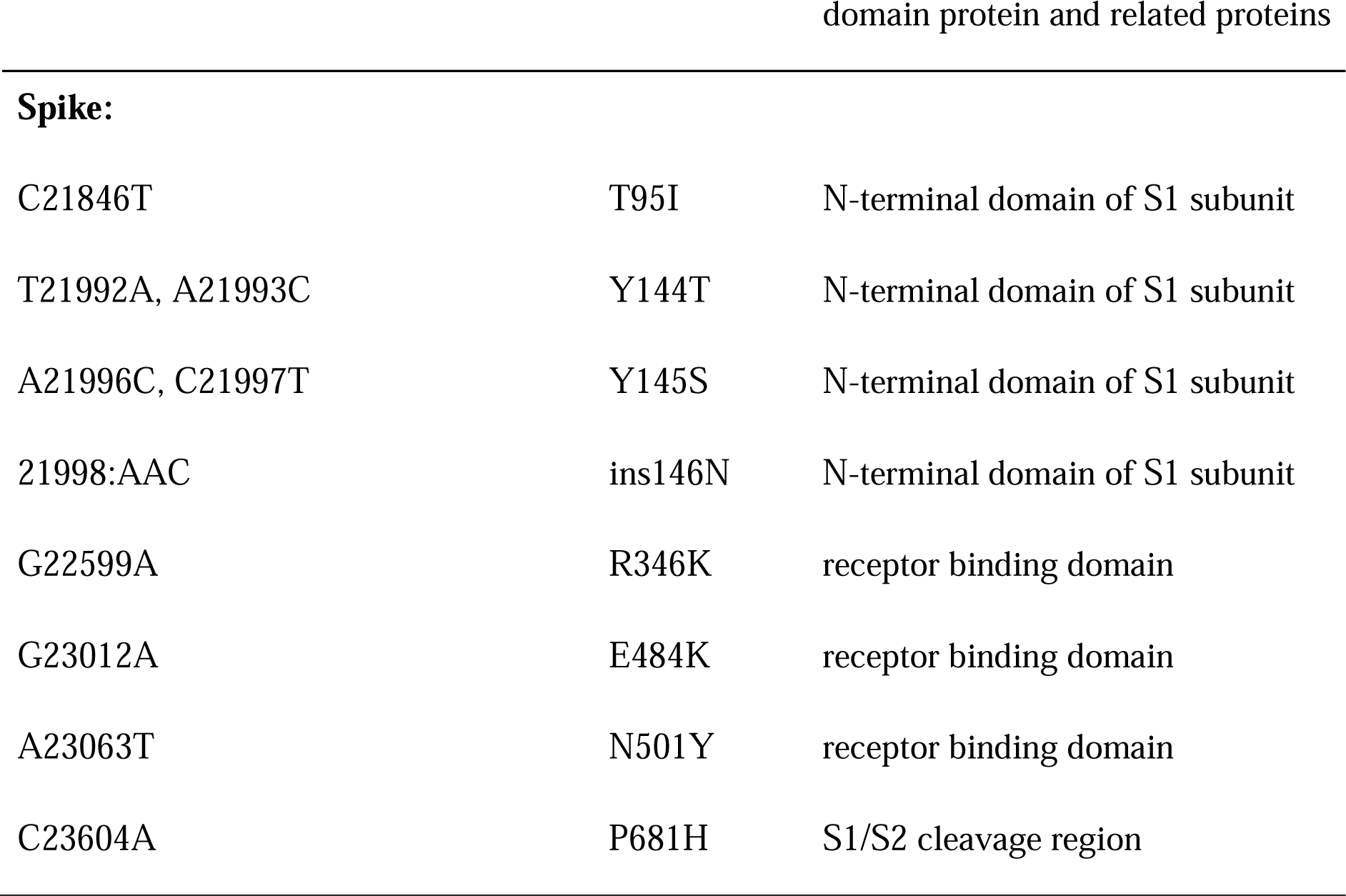
Nucleotide and amino acid substitution pattern of B.1.621

The close phylogenetic relationship of the SARS-CoV-2 sequences belonging to the B.1.621 lineage with other sequences from representative lineages circulating worldwide and those circulating in Colombia suggested a recent origin from the parental lineage B.1 (Figure 2a), which was corroborated through the lineage designation (https://github.com/cov-lineages/pango-designation/issues/57) (supplementary table 2). B.1.621 lineage has recently spread to fourteen departments, with a major representation in the Caribbean region of Colombia (Figure 2b) (https://microreact.org/project/5CAiK3qCMaEgE4vYkKVpZW/b7113efc). No recombination events were found throughout the whole genome (data not shown). At least 9 codons in the Spike protein displayed a signal suggestive of positive selection (supplementary table 3).

**Figure 2.**
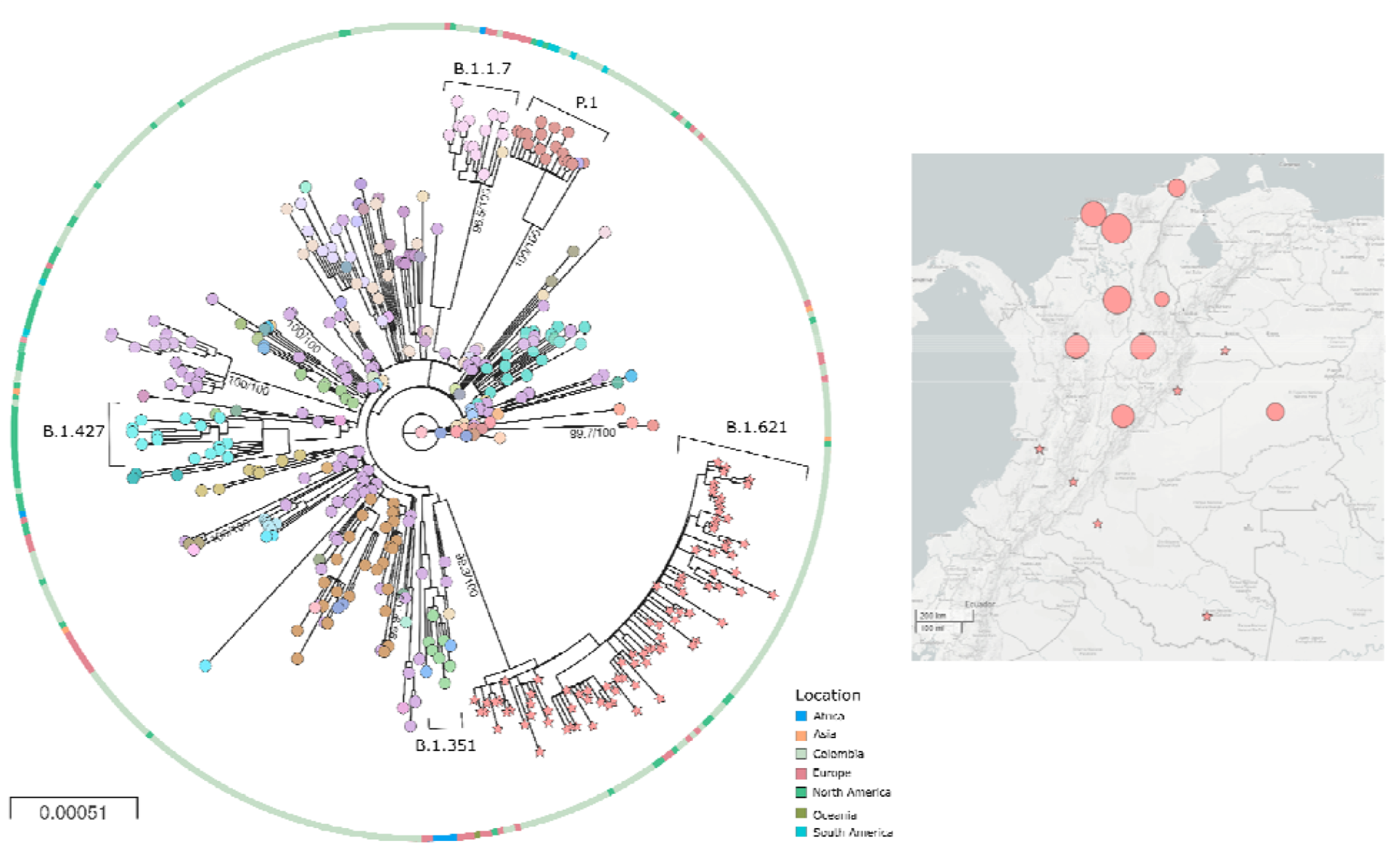
Phylogeny and distribution of SARS-CoV-2 b.1.621 variant in Colombia A) Phylogenetic tree of the new lineage of SARS-CoV-2 emerging from B.1.621 lineage (pink stars). The tree was reconstructed by maximum likelihood with the estimated GTR+F+I+G4 nucleotide substitution model for the dataset of 434 genomes. The interactive tree can be accessed in the following link: https://microreact.org/project/5CAiK3qCMaEgE4vYkKVpZW/b7113efc. B) Map of distribution of lineages across the country.

## 4. Discussion

All genomes belonging to the B.1.621 lineage available until the end of April 2021 were included in this study, with the earliest collection date on January 11th. 2021, corresponding to a sample collected in the department of Magdalena, Colombia (EPI_ISL_1220045). While a very high and unexplained genetic distance is found between every B.1.621 sequence and all closely related sequences of the parental B.1 lineage, the whole branching pattern and intra-lineage distance suggest low diversification that can be explained by its recent origin. The spread in the country early during the third peak of the pandemic could be explained by a combination of factors, including social exhaustion as well as the genetic background of the emerging lineage, leading to changes in transmission.

In Colombia, the current strategy for SARS-CoV-2 genomic surveillance includes sampling in principal and border cities, in special groups of interest, in patients with distinctive clinical features and severity, and finally in community transmission with an unusual increase in cases (https://www.ins.gov.co/Noticias/Paginas/coronavirus-genoma.aspx). The high frequency observed of the emerging B.1.621 lineage also could be related to the strengthening of the SARS-COV-2 genomic surveillance during the third peak of the pandemic in Colombia through the implementation of the National Laboratory Network for SARS-CoV-2 sequencing.

With this approach, we expect to characterize an approximate 1% of the cases and determine the adjusted frequency of the lineage in the country and to evaluate the possible predominance and the replacement of other lineages in the country. For this, intensified genomic characterization will be carried out with a multi-stage sample design throughout the national territory.

Since the last trimester of 2020, several convergent substitutions have been evidenced in the lineages of SARS-CoV-2 explained by a high rate of genetic variability by wide naive population, the selection pressure by monoclonal antibody therapies (Liu et al., 2021; Wibmer et al., 2021) and vaccination (Garcia-Beltran et al., 2021; Wang et al., 2021). Substitutions in the spike protein are common, however some distinctive substitutions have relevant characteristics for instance, the presence of E484K has been associated with lower neutralizing activity from convalescent plasma (Liu et al., 2021). The 69/70 deletion spike together with the E484K and N501Y substitutions decrease the ability to neutralize antibodies (Xie et al., 2021). Conversely, the P681H substitution in the S1/S2 furin cleavage site, although increase spike cleavage by furin-like proteases does not significantly impact viral entry or cell-cell spread in vitro (Örd et al., 2020) neither with higher infection rate or higher prevalence.

The insertion, 145N in the spike protein of B.1.621 is the first evidenced in this position in SARS-CoV-2, this insertion could affect the S1 closed-open conformation and the subsequent binding to the ACE2 (Berger and Schaffitzel, 2020), however its implications in terms of infection, transmission and pathogenesis are still unknown.

Although B.1.621 does not meet all of the VOC classification criteria so far, the set of mutations gathered the Spike protein could confer a synergistic impact on attributes such as reduction of vaccine-induced protection from severe disease, increased transmission and disease severity (Centers for Disease Control and Prevention, 2021).

Thus, laboratory characterization and enhanced routine genomic epidemiology studies are still required in order to monitor the possible change of status of this variant, or the emergence/introduction of new SARS-CoV-2 variants in the country.

Finally, the B.1.621 lineage has nucleotide substitutions already included in the VOC real-time reverse transcription PCR (real-time RT-PCR) screening tests (Bal et al., 2021). This should be considered in the analysis of these screening tests because the occurrence of these substitutions in lineages not regarded as VOC could lead to overestimating the number of VOC cases.

The B.1.621 lineage has been identified so far in Colombia, USA, Spain, Netherlands, Denmark, Mexico, Germany and Curacao. The study was limited to genomic and evolutionary characterization. The public health implications must be to assess through the biological and epidemiologic roles.

## Data Availability

All data is available in GISAID database, the phylogenetic analysis is available in a microreact site https://microreact.org/project/5CAiK3qCMaEgE4vYkKVpZW/b7113efc.

https://www.GISAID.org

## Funding

This work was funded by the Project CEMIN-4-2020 Instituto Nacional de Salud. The funders had no role in study design, data collection and analysis, decision to publish, or preparation of the manuscript.

## Disclosure statement

No conflict of interest was reported by the authors.

## Acknowledgements

The authors thank the National Laboratory Network for routine virologic surveillance of SARS-CoV-2 in Colombia. We also thank all researchers who deposited genomes in GISAID’s EpiCoV Database contributing to genomic diversity and phylogenetic relationship of SARS-CoV-2. We thank Rotary International and Charlie Rut Castro for equipment’s donation. Finally, we thank red RENATA and Universidad Industrial de Santander for the workstation bioinformatic support.

## Data deposition

SARS-CoV-2 Colombian sequences belonging to the B.1.621 were deposited in GISAID under accession numbers: EPI_ISL_1220045, EPI_ISL_1582980, EPI_ISL_1424054, EPI_ISL_1424056, EPI_ISL_1582978, EPI_ISL_1820926, EPI_ISL_1424055, EPI_ISL_1582993, EPI_ISL_1582979, EPI_ISL_1424057, EPI_ISL_1820929, EPI_ISL_1820930, EPI_ISL_1820932, EPI_ISL_1820927, EPI_ISL_1424058, EPI_ISL_1582984, EPI_ISL_1582986, EPI_ISL_1582991, EPI_ISL_1820958, EPI_ISL_1582990, EPI_ISL_1582992, EPI_ISL_1582981, EPI_ISL_1582994, EPI_ISL_1582988, EPI_ISL_1820959, EPI_ISL_1820928, EPI_ISL_1582996, EPI_ISL_1632530, EPI_ISL_1582989, EPI_ISL_1820934, EPI_ISL_1582987, EPI_ISL_1820955, EPI_ISL_1820935, EPI_ISL_1582997, EPI_ISL_1820925, EPI_ISL_1820950, EPI_ISL_1820949, EPI_ISL_1820944, EPI_ISL_1820947, EPI_ISL_1820943, EPI_ISL_1820946, EPI_ISL_1820954, EPI_ISL_1821882, EPI_ISL_1820939, EPI_ISL_1820942, EPI_ISL_1820960, EPI_ISL_1820936, EPI_ISL_1820948, EPI_ISL_1820931, EPI_ISL_1582977, EPI_ISL_1820965, EPI_ISL_1821070, EPI_ISL_1821075, EPI_ISL_1821063, EPI_ISL_1820967, EPI_ISL_1820968, EPI_ISL_1821062, EPI_ISL_1821064, EPI_ISL_1821069, EPI_ISL_1821073, EPI_ISL_1821074, EPI_ISL_1821076, EPI_ISL_1821077, EPI_ISL_1821071, EPI_ISL_1821072, EPI_ISL_1820962, EPI_ISL_1821065, EPI_ISL_1821066, EPI_ISL_1821067, EPI_ISL_1821068, EPI_ISL_1824702, EPI_ISL_1824703, EPI_ISL_1824706, EPI_ISL_1824711, EPI_ISL_1824712, EPI_ISL_1824713, EPI_ISL_1824714.

## Footnotes

1 T: Threonine, Y: Tyrosine, S: Serine, N: Asparagine, R: Arginine, K: Lysine, E: Glutamic acid, P: Proline, H: Histidine

## Notes

### Competing Interest Statement

The authors have declared no competing interest.

### Author Declarations

Good morning, dear reviewers, the ethical considerations were clarified in the body of manuscript because it was approved by the ethics committee of the National Institute of Health of Colombia within the framework of the Cemin 4 project. The circulation of the new variant of SARS-CoV-2 has been disclosed nationwide with an official statement (attached) because my Institution is the national authority on public health. We are also waiting for the lineage assignment by the PANGOLIN tool: https://github.com/cov-lineages/pango-designation/issues/23 I kindly require the acceptance of the preprint as soon as possible for its scientific divulgation that includes genetic characterization. Thank you very much for your prompt response. Regards

