## Supplemental Table 3 for "Characterization of the emerging B.1.621 variant of interest of SARS-CoV-2" (from another version of this preprint: version 1, 2021-05-10)

**Table 3. Codons under Positive selection of SARS-CoV-2 genome**

| FEL (Fixed Effects Likelihood) | | | | | | |
|---|---|---|---|---|---|---|
| **Gene** | **Codon** | **α** | **β** | **LTR** | **p-value** | |
| **ORF1ab** | 3729 | 0 | 40.049 | 1.27 | 0.2598 | |
| **ORF3a** | 57 | 0 | 18.46 | 1.686 | 0.1941 | |
| **ORF8** | 11 | 0 | 10.082 | 1.121 | 0.2896 | |
| **ORF8** | 67 | 0 | 10.518 | 3.472 | 0.0624 | |
| **S** | 95 | 0 | 7.050 | 2.300 | 0.1294 | |
| **S** | 144 | 0 | 42.400 | 10.388 | 0.0013 | |
| **S** | 484 | 0 | 28.495 | 3.087 | 0.0789 | |
| **S** | 501 | 0 | 7.878 | 1.840 | 0.1750 | |
| **S** | 681 | 0 | 7.853 | 2.8 | 0.0942 | |
| **MEME (Mixed Effects Model of Evolution)** | | | | | | |
| **Gene** | **Codon** | **α** | **β+** | **Pr[β=β+]** | **LRT** | **p-value** |
| **ORF1ab** | 3729 | 0 | 146.327 | 2.81E-01 | 1.207 | 0.2821 |
| **S** | 144 | 0 | 163.83 | 407 E-01 | 830 | 0.0089 |
| **S** | 501 | 0 | 45.983 | 243 E-01 | 1.135 | 0.2939 |
| **S** | 681 | 0 | 21.591 | 601 E-01 | 2.122 | 0.1704 |
